# Scleroderma Clinical Trials Consortium Classification Criteria for Systemic Sclerosis Heart Involvement

**DOI:** 10.1101/2025.11.11.25339972

**Authors:** Laura Ross, Andrew T. Burns, André La Gerche, Dylan Hansen, J. Gerry Coghlan, Wendy Stevens, David Prior, Alan Pham, Penny McKelvie, Chiara Bellocchi, Yolanda Braun Moscovici, Cosimo Bruni, Patricia Carreira, Tracy Frech, Sabrina Hoa, Marie Hudson, Vivien Hsu, Andrea Hsiu Ling Low, Marco Matucci-Cerinic, Benjamin Medina Fonseca, Sue-Ann Ng, Tatiana Rodriguez Reyna, Joanne Sahhar, Mohamed Talaat, Susanna Proudman, Alessandra Vacca, Murray Baron, Mandana Nikpour, the SCTC Cardiac Working Group

## Abstract

**Objectives:** Systemic sclerosis (SSc) heart involvement (SHI) is an enigmatic disease manifestation associated with high mortality. The Scleroderma Clinical Trials Consortium (SCTC) Cardiac Working Group developed SHI classification criteria to enable systematic investigation of this condition.

**Methods:** An international, inter-disciplinary working group was assembled. Using consensus methods and existing literature, provisional SHI classification criteria items were developed. Continuous consensus exercises and a discrete choice experiment were performed to reduce items and derive individual item weights. The sensitivity and specificity of the classification criteria were tested in an independent cohort (n=168) of SHI (cases) and non-SSc heart disease (controls).

**Results:** The working group agreed that the SCTC SHI Classification Criteria should identify the direct effects of SSc on the heart and exclude the complications of other SSc manifestations or cardiac co-morbidities. The final classification criteria include 23 items measuring cardiac fibrosis, inflammation, arrhythmias and small vessel vasculopathy. No single item is pathognomonic for SHI, with a requirement for the presence of abnormalities across multiple histopathological, imaging, serological and, or clinical domains to be present to secure a diagnosis. A classification criteria score of ≥11 identified SHI with a sensitivity of 78% and specificity of 96%, with an area under the curve of 0.87 (0.80-0.93). This threshold correctly identified >90% of cases of SHI.

**Conclusion:** The newly derived SCTC SHI Classification Criteria have high sensitivity and specificity for SHI. Application of these criteria will enable standardised classification of patients in studies to facilitate future investigation of this important disease manifestation.

**Graphical abstract:** *Abbreviations:* AUC: area under the curve; IHD: ischaemic heart disease; PAH: pulmonary arterial hypertension; SHI: systemic sclerosis heart involvement; SRC: scleroderma renal crisis
*Alt text:* Flow chart representing the steps to define systemic sclerosis heart involvement criteria, staring from scope and planning, item generation, item reduction and weighting and concluding with defining a classification threshold and testing performance of criteria. A criteria score of 11 or greater classifies a patient as having systemic sclerosis heart involvement.

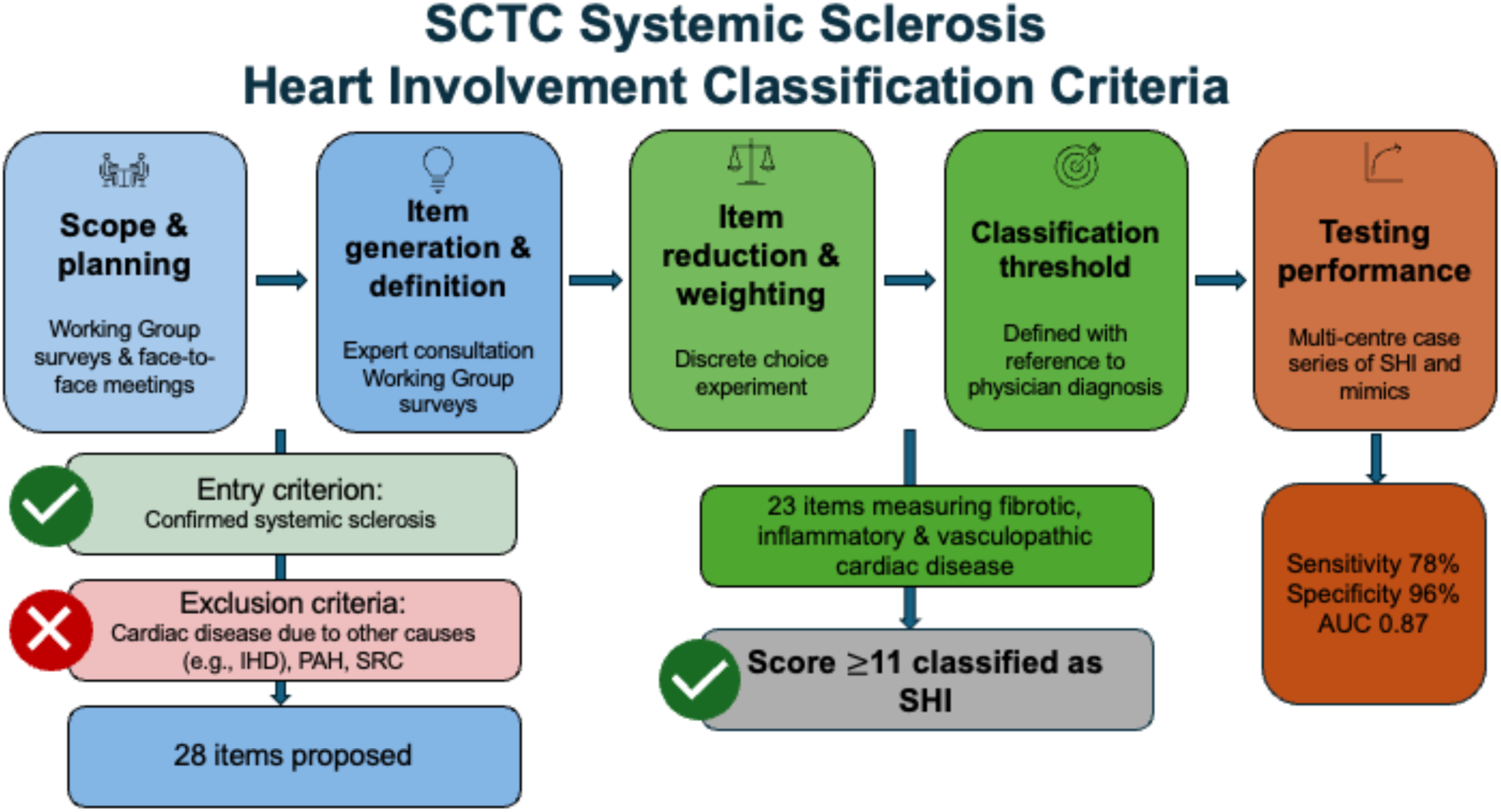

**Key messages:**

- This study presents the first classification criteria for the identification of systemic sclerosis heart involvement.
- A score of ≥11 identifies systemic sclerosis heart involvement with high sensitivity and specificity.
- Standardised criteria enable identification of biomarkers and risk predictions models and lead to effective treatments for heart involvement.

## Introduction

Systemic sclerosis (SSc) can affect all major organ systems, including the heart, with a spectrum of cardiac disease ranging from florid myocarditis to asymptomatic chronic pericardial effusion.[1] Studies since the 1940s have reported the direct effects of SSc on the heart,[2] and autopsy studies document heart involvement in up to 90% of cases.[3–5] SSc-associated heart involvement (SHI) remains a leading cause of mortality with no disease-modifying therapy available.[1, 6, 7]

A consensus definition of SHI has been proposed, stating that SHI is defined by the presence of cardiac abnormalities predominantly attributable to SSc.[8] The definition requires SHI to be confirmed by cardiac investigation,[8] with increasing evidence demonstrating the importance of even subtle changes of cardiac structure and function.[9–12] There remains an absence of validated criteria by which to identify SHI. Highly variable case ascertainment means the burden of SHI is variably quantified, the natural history of SHI is poorly defined and there are limited methods to identify high-risk patients.[1]

In response, the Scleroderma Clinical Trials Consortium (SCTC) Cardiac Working Group sought to develop SHI classification criteria. These criteria would enable accurate identification and quantification of the burden of SHI, identification of important risk and prognostic factors for SHI and facilitate therapeutic trials.

## Methods

### Investigators

The SCTC Cardiac Working Group included 24 rheumatologists, 5 cardiologists and 2 anatomical pathologists. The American College of Rheumatology propose methodological guidance for the development of classification criteria (https://rheumatology.org/criteria). These methods were adapted to develop the SHI classification criteria (Figure 1).

**Figure 1:**
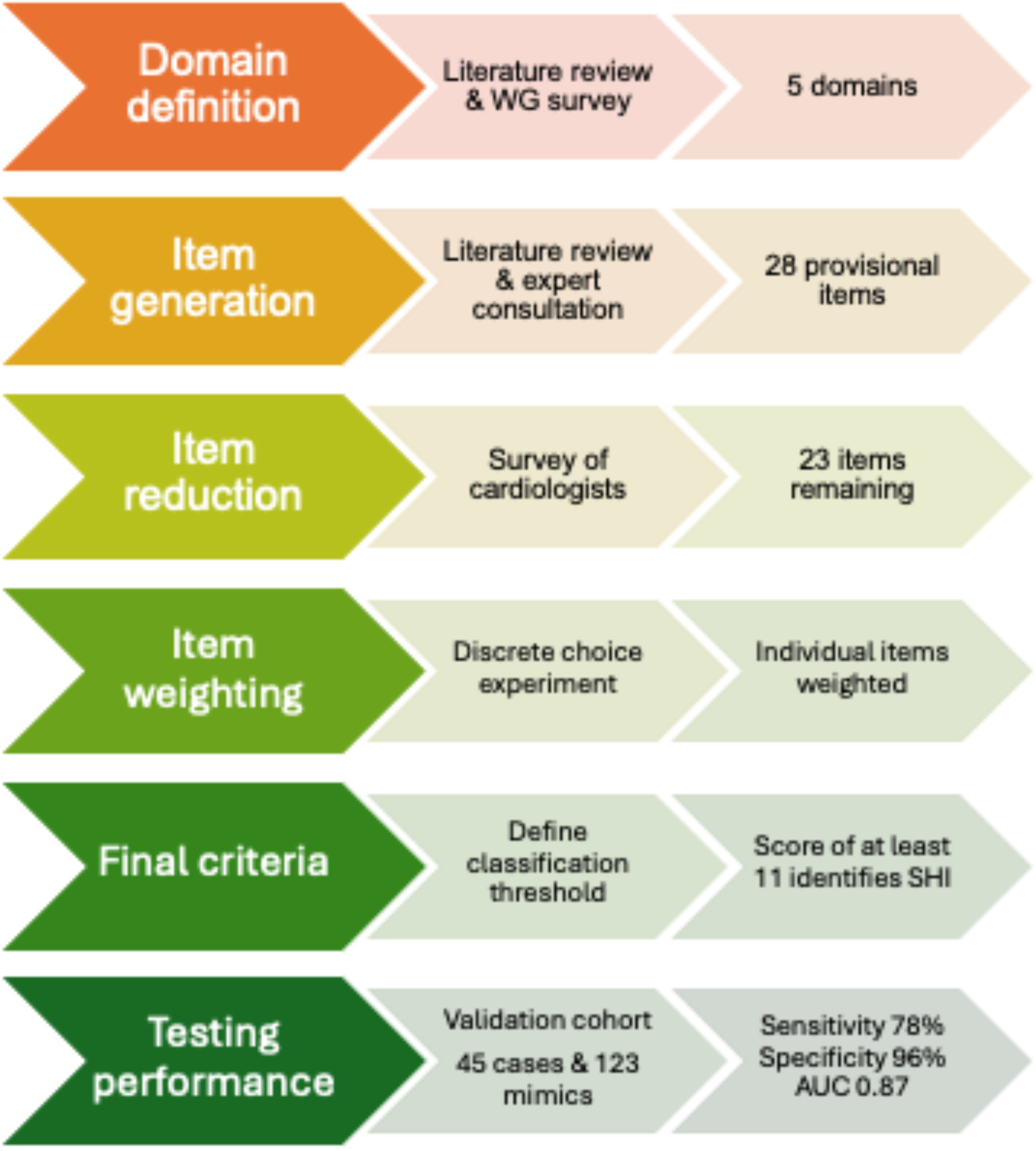
Development of the SCTC Systemic Sclerosis Heart Involvement Classification Criteria. *Abbreviations:* AUC: area under the curve; WG: working group *Alt text:* Flow chart summarising methodological process of development SCTC SHI Classification criteria, including domain definition, item generation, item reduction and weighting and testing the performance of the final criteria showing high sensitivity (78%) and specificity (96%) for the identification of systemic sclerosis heart involvement.

### Domain definition

A scoping literature review was performed to identify previous definitions of SHI and generate potential classification criteria items.[1] Using these results, a survey to define the domains of disease to be included in the SHI classification criteria was developed. Working group members completed a single-round survey, with results presented at an in-person meeting. An *a priori* threshold of 70% agreement for inclusion or exclusion of a domain was set.

### Expert consultation

Rheumatologists (n=4), cardiologists (n=5) and anatomical pathologists (n=2) were consulted to generate provisional classification criteria items. Cardiologists included had expertise in cardiac imaging (echocardiography and cardiac magnetic resonance imaging (CMR)), heart failure and pulmonary vascular disease. Anatomical pathologists had subspecialty expertise in the interpretation of cardiac and skeletal muscle histopathology. The expert panel was presented with results of the literature review and working group survey and were asked to suggest items to measure each domain. In an iterative process, experts were asked to provide feedback on the items suggested by other members of the expert panel. Disagreement about proposed items was resolved by consensus. A face-to-face meeting including two cardiologists and two rheumatologists from the expert panel reviewed all suggested items, and provisional items were grouped according to the underlying pathophysiological mechanisms that contribute to SHI: myocarditis, myocardial fibrosis, vascular abnormalities and pericardial abnormalities. All preliminary items were then distributed to members of the expert panel to confirm agreement with the inclusion and grouping of provisional items.

### Item reduction

A group of 15 cardiologists from Europe, United Kingdom (UK), North America and the Asia-Pacific, with experience in the assessment and management of heart disease due to SSc and other autoimmune diseases were invited to participate in an online survey. Respondents were asked to rate the specificity of each provisional criterion for SHI on a Likert scale from (1) highly unlikely to indicate SHI to (7) highly likely to indicate SHI. Items that were rated as at least somewhat likely (score ≥5) by at least 50% of experts were retained for the next phase. Survey results were presented to all participating cardiologists who were asked to assess the face and content validity of the proposed list of classification items.

### Item weighting

Cardiologists and rheumatologists were invited to participate in an online discrete choice experiment to assign weights to individual items using multicriteria decision analysis.[13] Respondents were presented with a series of paired scenarios and asked to rate which of the scenarios was most likely a case of SHI. The discrete choice experiment was hosted on the 1000minds platform (http://www.1000minds.com). The PAPRIKA method was applied to derive a relative ranking for each provisional classification criteria item.[13] Item weights were calculated by rounding the relative item weights to the nearest integer.

### Defining a classification threshold

Twenty cases, representing a range of SHI presentations and heart disease mimickers were collected to enable definition of a classification threshold of SHI. Cases were selected to include the spectrum of features reported to indicate the presence of SHI.[1] These cases were presented to cardiologists and rheumatologists in a standardised format.

All cardiologists who completed the item reduction survey and 22 rheumatologists from Europe, UK, North and Central America and the Asia-Pacific were asked to rate their diagnostic certainty about each case on a numerical rating scale from 0 to 20, where 0 = definitely not SHI to 20 = definitely SHI. Respondents were not provided with data from previous phases of the criteria development and were blinded to other participants’ rankings. Median scores for each case were calculated by pooling all participant responses.

Physician determined presence of SHI was the gold standard comparison to define the SHI classification threshold. This is consistent with the current SHI diagnostic standard of physician diagnosis, supported by cardiac investigations and exclusion of other cardiac disease.[8] This approach has been applied when developing classification criteria for other systemic autoimmune diseases.[14]

Analysis of case ranking scores showed a score of 15 was the threshold above which a case was ranked in the top quartile of cases most likely to be SHI. This was defined as the threshold for a case to be considered highly likely or definite SHI. This threshold was set to optimise the specificity of the classification criteria given their intended purpose for use as a research tool. Cases that were ranked by >50% of experts as highly likely SHI (score ≥15) were considered positive for SHI and were used to define the criteria threshold for identifying SHI.

Each of the 20 cases was assigned a SHI classification criteria score by summing the individual item weights derived from the discrete choice experiment. Cases that scored around the provisional classification threshold were reviewed again by two expert rheumatologists and one cardiologist to ascertain which cases could be confidently classified as having SHI. A threshold for the classification of SHI was identified based on targets of >90% specificity and >80% sensitivity.

### Testing the performance of the classification criteria

SCTC members were invited to submit cases of SHI and non-SSc associated heart disease, representing the spectrum of heart disease observed in clinical practice to form a cohort of cases in which to test the new criteria. Cases of non-SHI were collected to form a control group. Submitted cases could be either historical or contemporary. No case used to define the classification threshold was included in this cohort. Cases were all submitted prior the definition of any classification thresholds for SHI. Submitting members did not participate in the item generation or reduction phases of the criteria development.

Data were entered into a standardised data collection form that captured clinical symptoms, examination findings, and electrocardiographic, imaging and histopathological findings. It was not a requirement that all investigations be present for a case to be included, as the goal was to collect a real-world cohort of SHI cases reflective of contributing members’ clinical experience. Contributing members were asked to nominate whether their patient had SHI or heart disease of another aetiology, using a Likert scale that ranged from (1) definitely SHI to (7) definitely not SHI. This clinical assessment served as the reference gold standard test for the presence of SHI. Physician ratings of cases were reviewed by two authors (LR, MB) to ensure consistency of rating of cases as confirmed SHI or not SHI before any calculation of classification criteria scoring. The clinical characteristics of those with and without SHI in the validation cohort were compared using chi-squared or Fisher’s exact test as appropriate for discrete variables and Wilcoxon rank-sum test for non-normally distributed continuous variables. A SHI classification criteria score was calculated for each submitted case. Cases that were positive for any of the SHI classification exclusion criteria were scored as 0. Any missing items were scored as 0, i.e. if case had no CMR results, the case would score zero points for any CMR classification criteria items.

A case was considered positive for SHI if the submitting physician rated the case as (1) Definitely SHI or (2) Highly likely SHI. All other cases of uncertain SHI or non-SSc heart disease were considered as negative for SHI. Performance of the classification criteria was assessed using a preliminary threshold of ≥11 points. This threshold was selected based on sensitivity and specificity testing. Further sensitivity analyses of alternate classification thresholds were tested. Sensitivity and specificity were calculated, with discrimination tested by calculating the area under the curve (AUC) of receiver operator characteristic curves. Logistic regression analysis was performed to calculate the odds ratio of a specific SHI classification criteria threshold detecting SHI. The percentage of correctly classified cases was recorded.

Study data were collected and managed using REDCap electronic data capture tools hosted at The University of Melbourne. All statistical analyses, excluding analysis of the discrete choice experiment results, were performed using STATA 18.0 (StataCorp, College Station, TX, USA). Ethics approval for this study was granted by the Human Research Ethics Committee at St Vincent’s Hospital Melbourne (LRR 195/20).

## Results

### Domain generation

The Working Group reached consensus to develop classification criteria for SHI that could be applied to clinical trials and observational studies. The group agreed that classification criteria should focus on primary heart involvement and not include cardiac complications of other SSc manifestations such as pulmonary arterial hypertension (PAH), interstitial lung disease (ILD) or renal crisis (SRC). It was agreed that SHI classification criteria should distinguish the direct effects of SSc on the heart from other cardiac co-morbidities.

Twenty-two working group members (15 rheumatologists and 7 cardiologists) were invited to participate in an initial survey to define the domains of cardiac disease to include in the classification criteria. Sixteen (73% response rate) complete responses were received. The following domains of cardiac disease were identified to be included in the SHI classification criteria (>70% agreement for inclusion): myocardial fibrosis, myocardial inflammation, arrhythmias, conduction abnormalities, pericardial abnormalities. Sixty-nine percent of respondents agreed perfusion abnormalities should be a domain. Valvular heart disease was omitted at this stage.

### Item generation

With consideration to the scoping literature review[1] and domain generation survey results, the expert panel iteratively generated potential items to measure each of the proposed SHI domains. The expert panel agreed that arrhythmias and conduction abnormalities are clinical manifestations of underlying myocardial fibrosis or inflammation, so items pertaining to these domains were grouped within the fibrosis and inflammation domains. Vascular abnormalities that may result from SHI were considered, given this domain almost reached the threshold of agreement. The expert panel agreed that microvasculopathy could be consistent with SHI, therefore, a vascular domain was retained.

In recognition that changes of SHI can mimic several other conditions, the expert panel agreed to have exclusion criteria as part of the SHI classification criteria. The following exclusion criteria were developed: (i) A diagnosis of SHI can only be made in the setting of an established diagnosis of SSc; (ii) SHI can only be diagnosed when other causes of cardiomyopathy, particularly infiltrative diseases, have been reasonably excluded. Other causes of cardiomyopathy to exclude are cardiac disease secondary to pulmonary vascular disease, ischaemic heart disease, hypertensive heart disease, cardiac disease secondary to renal disease, dilated cardiomyopathy (e.g. genetic, post non-SSc myocarditis, alcohol), cardiac sarcoidosis, cardiac amyloidosis, iron overload, and Fabry’s disease. The expert group generated 28 items to measure SHI across four domains of cardiac disease: myocardial fibrosis, myocardial inflammation, vascular abnormalities, and pericardial abnormalities.

### Item reduction and weighting

Fifteen cardiologists were invited to complete a survey rating the specificity of the preliminary criteria. Twelve (80%) complete responses were received. Three items (left ventricular hypertrophy, atrial fibrillation, cardiac tamponade) were omitted in this phase. Reduced ventricular ejection fraction and ventricular dysfunction were combined into a single item. Twenty-one rheumatology and cardiology experts participated in an online discrete choice experiment. A relative ranking and item weight of each SHI classification criteria item were generated. The final SHI classification criteria items are listed in Table 1.

**Table 1:**
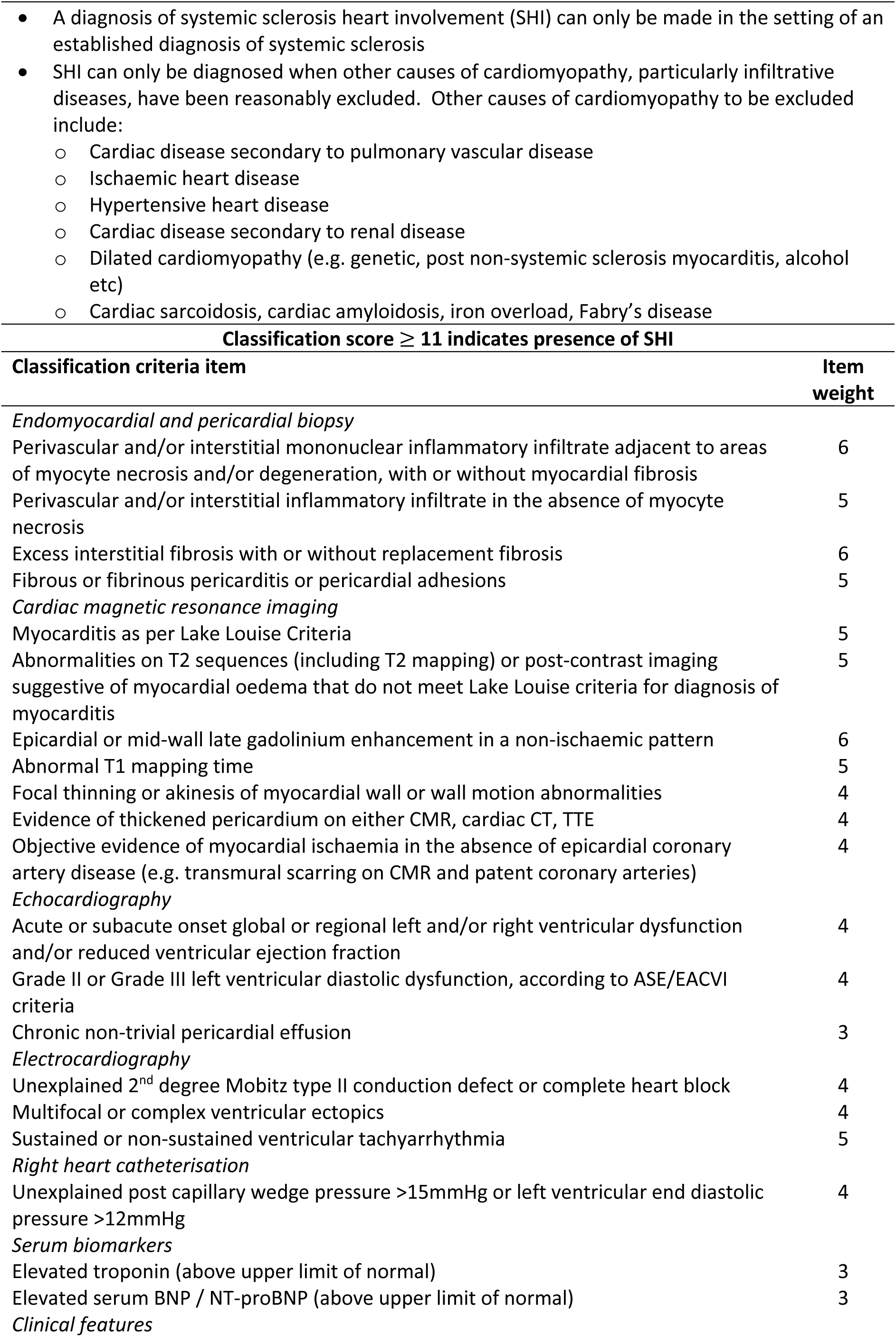
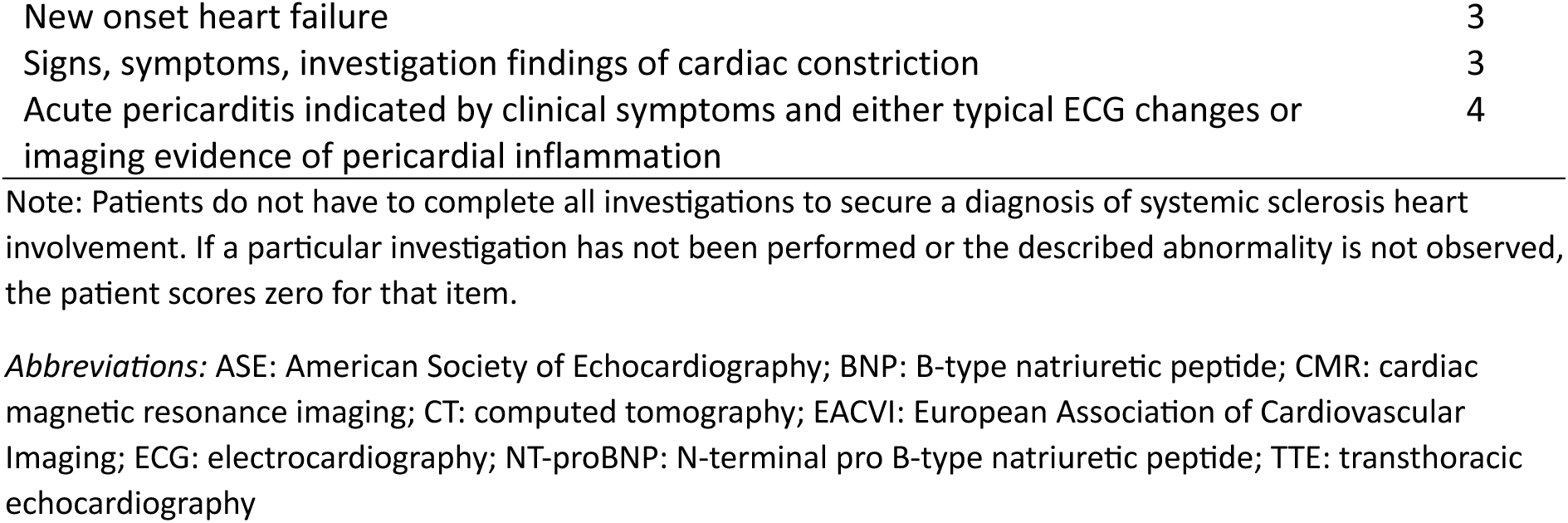
SCTC Systemic Sclerosis Heart Involvement Classification Criteria.

### Defining the classification threshold

Twenty-eight survey responses were received (82% response rate). Inspection of case rankings revealed a significant spread of results across the Likert scale. Seven (35%) cases had more than 50% of respondents rate the case with a score ≥15. These cases were considered as positive for SHI. Eleven (55%) cases had a median ranking of >10, equating to possible or likely SHI. A SHI classification score was calculated for each case by summing individual item weights. A preliminary classification threshold of ≥11 points identified cases considered to be SHI as determined by physician diagnosis with a sensitivity of 100% (95%CI 66.37-100%) and specificity of 81.82% (95% CI 48.22%-97.72%).

Table 2 describes the assembled test cohort. A total of 168 cases were submitted of which 45 (27%) were considered to have definite SHI. The median SHI classification score of the testing cohort was 3 (IQR 0-10). (Cases were scored zero if they met any of the SHI exclusion criteria.) Forty cases (24%) had a SHI criteria score ≥11 points. A classification criteria threshold of ≥11 points correctly classified 91.07% of this external testing cohort with a sensitivity and specificity of 77.78% and 95.93%, respectively. This threshold had good discrimination with an AUC of 0.87 (0.80-0.93) and a diagnostic odds ratio of 82.60 (95%CI 26.47-257.72, p<0.01). The sensitivity and specificity of other thresholds were tested (Supplementary Index 1). Owing to the emphasis placed on specificity for the development of classification criteria, a threshold of ≥11 was considered the most appropriate threshold as this cut point had an excellent AUC, high specificity and high odds ratio for the diagnosis of SHI.

**Table 2:**
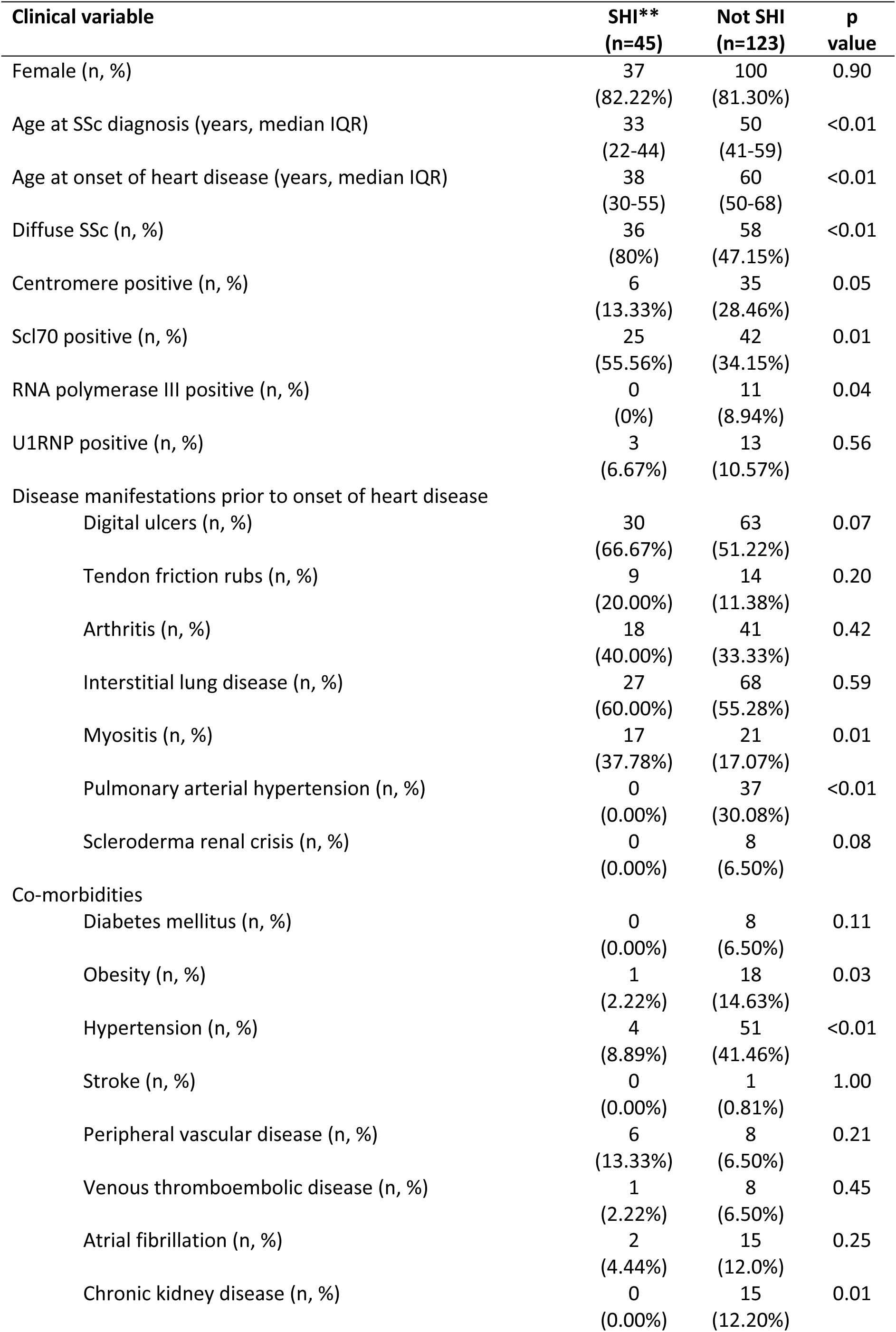
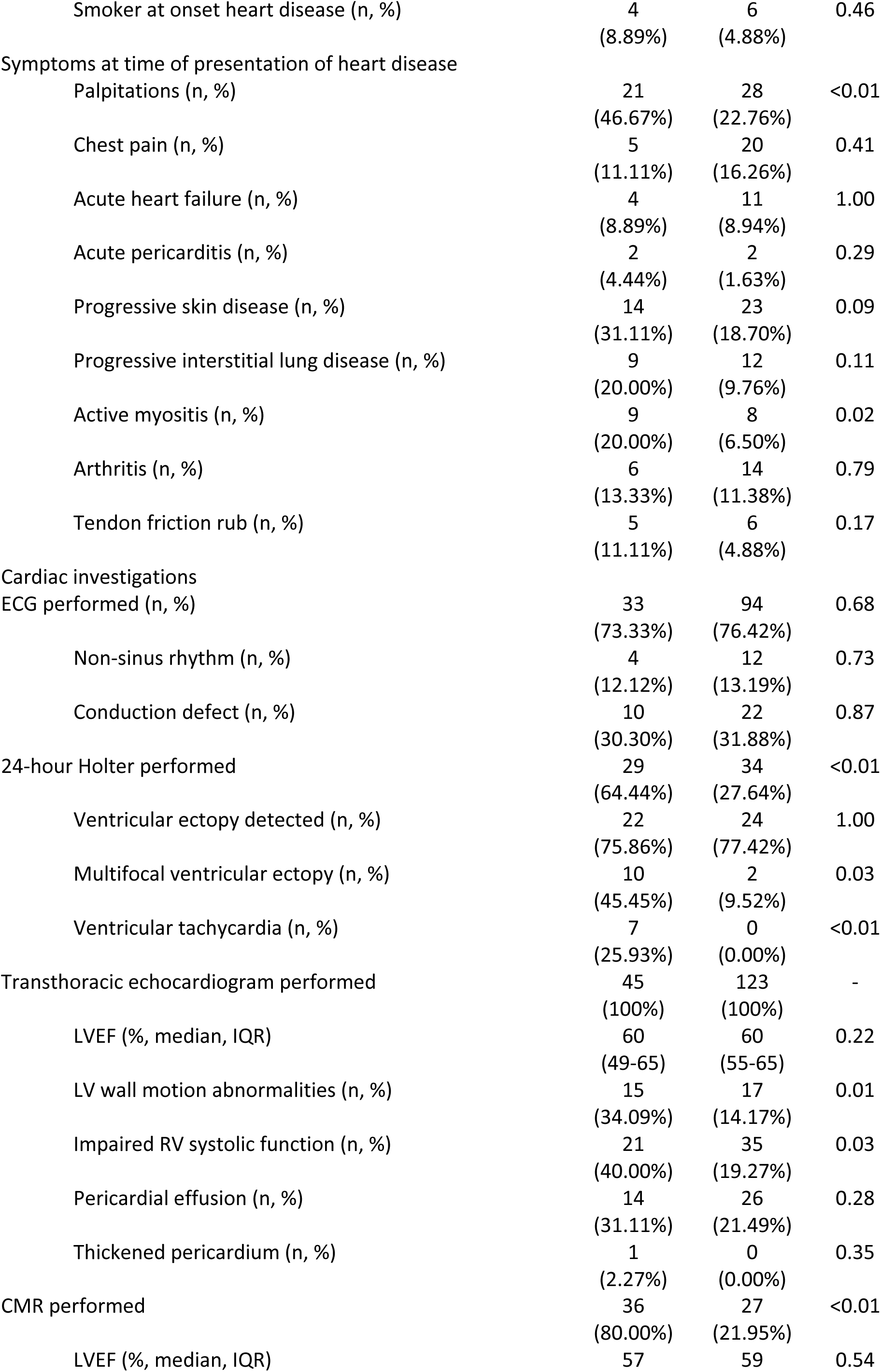
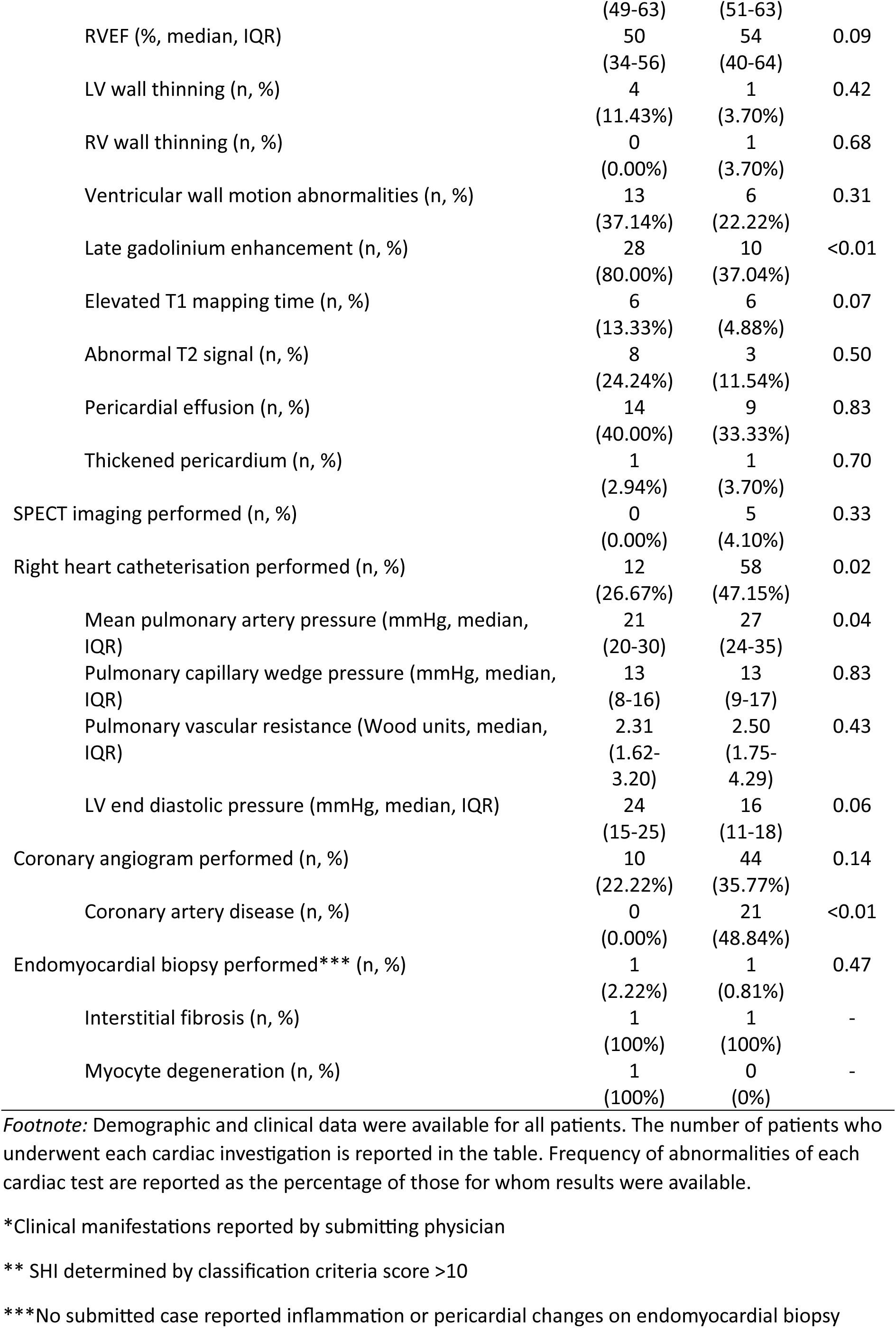
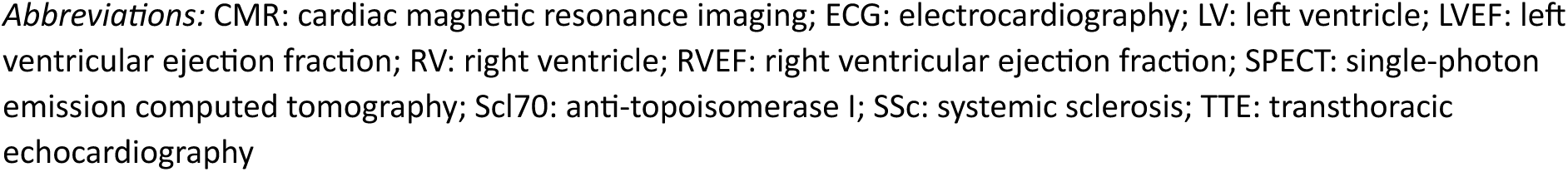
Clinical characteristics of the validation cohort.

## Discussion

Following a formal iterative process of testing, we propose the first classification criteria for SHI. Previous work has developed a definition of SHI,[8] and these are clinical parameters that can operationalise this definition. These criteria include both invasive and non-invasive measures of the hallmark histopathological lesions of SHI, namely inflammation, microvasculopathy and fibrosis.[4] The development of the criteria has intentionally targeted the measurement of these changes using investigation modalities currently applied in clinical practice.

Reflective of the diversity of potential SHI manifestations and the lack of a single pathognomonic feature of SHI,[1] no single criterion is sufficient for a patient to be classified as having SHI. However, the criteria are not an exhaustive list of all cardiac abnormalities that can be attributed to SSc. The criteria have been developed with an intentional preference for item specificity over sensitivity. For example, diastolic dysfunction is a common cardiac abnormality associated with SSc,[15, 16] but there is overlap between echocardiographic parameters of healthy individuals and those with diastolic dysfunction.[17] Normal ageing is associated with impaired ventricular relaxation.[17] Therefore, a higher threshold of positivity, Grade II diastolic dysfunction, was established for this item to capture changes more specific to underlying pathology rather than potentially normal phenomena.

Use of CMR is becoming more routine and it is the only non-invasive technique able characterise myocardial inflammation and scar.[18, 19] CMR parametric mapping techniques can identify diffuse myocardial oedema and fibrosis. CMR has been proposed as a screening tool for SHI,[20] and its use is likely to rapidly expand in the diagnosis of both sub-clinical and manifest cardiac disease in SSc. Endomyocardial biopsy (EMB) has utility in the diagnosis of SHI, however the procedure carries potential risks and may only be available at specialist centres. Additionally, it may not be routinely required with the increasing availability of CMR. Our study cohort reflected this with the infrequent use of EMB to secure a diagnosis of SHI. Furthermore, there is no universal histopathological criteria for diagnosis of EMBs.[21] Our criteria make reference to the Dallas criteria,[22] widely used histopathological criteria for the identification of myocarditis.[21] There are no routinely used pathological criteria to define excess fibrosis, with excess fibrosis being defined as that in excess of what would be expected for the patient’s age. Qualitative descriptions of excess fibrosis are included in the descriptions and classification of other cardiomyopathies.[23–25]

These classification criteria have included explicit exclusion criteria. In the absence of a single diagnostic test or pathognomonic finding of SHI, there remains a requirement to exclude other causes of cardiac disease.[8] For example, co-morbid PAH and SRC are exclusion criteria because of their known cardiac sequelae. It is not currently possible to distinguish between cardiac complications of PAH and SRC from SHI. As understanding of the genetic and molecular underpinnings of SHI improve, it may be possible to distinguish between primary and secondary cardiac consequences of SSc, potentially removing the need to have exclusion criteria as part of any SHI classification criteria.

These criteria will not capture all patients who have SHI. They have been intentionally designed to have the highest possible specificity, to minimise the identification of false-positive cases.[26] Classification criteria are intended for use in clinical research rather than clinical practice. A failure to meet these criteria should not preclude a patient from being treated for SHI. A high clinical suspicion of SHI but a failure to meet the classification threshold may be more likely in cases where no CMR or EMB can be obtained. These criteria can guide clinicians as to the expected cardiac manifestations that are part of the spectrum of SHI and suggest an investigation framework. However, these criteria do not recommend that all cardiac investigations be performed in all SSc patients. Nor should the list of exclusion criteria be interpreted as a list of tests that must be obtained in all cases of suspected SHI.

This study has several strengths, including the use of expert consensus and decision analysis to derive and weight the SHI classification criteria items. Input from experts in the fields of rheumatology, cardiology, cardiac imaging and anatomical pathology was sought. There was wide geographic representation from experts and test cases from across North and Central America, Europe and the UK, Asia and Australia. This was intentional to develop criteria that can be widely applied. The criteria were tested in a patient cohort developed without knowledge of the SHI classification criteria, minimising circularity of reasoning in the testing of the criteria.

There are limitations of this study. No case had results for all cardiac investigations that can be used to identify SHI. The testing of these criteria was performed in a cohort reflective of real-world practice. It is likely that some cases rated as somewhat likely or uncertain by the submitting physician may have had a diagnosis of SHI secured or definitively excluded if additional investigations had been performed. Many cases were retrospective cases from contributing physicians’ clinical practice, meaning cases may be subject to the recall bias of contributing physicians. Our overall test cohort was small. Florid SHI is an uncommon manifestation of a rare disease, with a prevalence of approximately 5% in large SSc cohorts.[27, 28] Therefore, the generation of a combined cohort of 168 patients from multiple centres globally is one of the most significant efforts to collate detailed data about the various cardiac manifestations of SSc. These criteria would benefit from prospective evaluation in larger cohorts that include greater numbers of patients with SHI to further interrogate their discrimination between SHI and non-SHI heart disease.

Testing and validating classification criteria are challenging in the absence of a gold standard more robust than that of the physician diagnosis. A physician diagnosis of SHI is the current reference standard for SHI with no minimum test requirement to secure the diagnosis. Such a test is subject to significant inter-observer variability. Even amongst experts we observed significant variation in the assessment of standardised cases. The expert review of controversial cases to reach a consensus opinion as to the presence of SHI is a strength of this project. However, the sensitivity and specificity of the final criteria may have been affected by the variability in physician assessment, although this is an inherent limitation of the first iteration of any classification criteria.

As methods of assessing cardiac inflammation, fibrosis and vasculopathy improve,[29–33] these criteria will need to be revisited, with re-consideration of both item definition and individual item weights. The classification threshold required to identify cases of definite SHI may change as newer investigation modalities are implemented in clinical practice.

In conclusion, we present the first classification criteria for SHI. Using a combination of consensus and data-driven approaches, we have developed criteria that have good sensitivity and excellent specificity and can identify SHI across the spectrum of disease. They represent a significant advance in this evolving field. We encourage their implementation in future studies to further validate their use and to encourage investigation of this important disease manifestation.

## Supporting information

Supplementary Index 1

## Acknowledgements

Study authors wish to acknowledge the SCTC Cardiac Working Group Members and both the cardiology and rheumatology experts who contributed responses to study surveys: Catherine Abric, Monica Alouian, Shervin Assassi, Murray Baron, Cosimo Bruni, Maya Buch, Andreea Bujor, Andrew Burns, Lorinda Chung, Benedict Costello, M.E. Csuka, Francesco Del Galdo, Christopher P Denton, Jeska de Vries-Bouwstra, Girish Dwivedi, Tracy Frech, J. Gerry Coghlan, Clive Handler, Ariane Herrick, Monique Hinchcliff, Alicia Hinze, Sabrina Hoa, Vivien Hsu, Marie Hudson, Laura Hummers, Jacqueline Joza, Kusano Kengo, Dinesh Khanna, André La Gerche, David Langleben, Thomas Medsger, Edward Miller, Roberta Montisci, Mandana Nikpour, Gene-Siew Ngian, John D Pauling, David Prior, Susanna Proudman, Tatiana Rodrigeuz-Reyna, Lawrence Rudski, Joanne Sahhar, Flora Sam, James Seibold, Virginia Steen, Wendy Stevens, Alessandra Vacca, Jaap van Laar; and Alison Hendry for methodological advice.

## Conflict of interest statement

LR: No conflict of interest to declare.

AB: No conflict of interest to declare.

ALG: No conflict of interest to declare.

DH: No conflict of interest to declare.

JGC: Research grants from Johnson & Johnson unrelated to submitted work. Speaker and consultancy fees from Johnson & Johnson and MSD.

WS: Research support: Janssen, Bristol Myers Squibb, Boehringer Ingelheim. Consultancy fees from Janssen, Certa, MSD, Boehringer Ingelheim. Honoraria from Janssen, GlaxoSmithKline, Boehringer Ingelheim

DP: No conflict of interest to declare.

AP: No conflict of interest to declare.

PM: No conflict of interest to declare.

CBe: No conflict of interest to declare.

YBM: No conflict of interest to declare.

CBr: Consultancy fees: Boehringer Ingelheim

PC: No conflict of interest to declare.

TF: No conflict of interest to declare.

SH: No conflict of interest to declare.

MH: No conflict of interest to declare.

VH: Research support from GSK and Novartis.

BMF: No conflict of interest to declare.

AHLL: Research support: Boehringer Ingelheim;

Consultancy fees: Janssen, Boehringer Ingelheim, Novartis;

Honoraria: Boehringer Ingelheim, Novartis

MMC: No conflict of interest to declare.

SN: No conflict of interest to declare.

TRR: No conflict of interest to declare.

JS: Research support: Janssen, Boehringer Ingelheim. Honoraria from Boehringer Ingelheim.

MT: No conflict of interest to declare.

SP: Research support: Janssen, Boehringer Ingelheim. Honoraria from Janssen, Boehringer Ingelheim, MSD.

AV: No conflict of interest to declare.

MB: No conflict of interest to declare.

MN: Research support: Janssen, BMS, GSK, AstraZeneca, Boehringer Ingelheim, UCB. Consultancy: Janssen, Certa, AstraZeneca, Boehringer Ingelheim, Eli Lilly

Honoraria: Janssen, GSK, AstraZeneca, Boehringer Ingelheim, Pfizer, UCB.

## Data availability statement

Data is available from corresponding author upon reasonable request.

