## Supplementary Index 1 for "Scleroderma Clinical Trials Consortium Classification Criteria for Systemic Sclerosis Heart Involvement"

### Supplementary material

*Supplementary Index 1: Sensitivity and specificity of tested classification thresholds for SHI*

| Classification Threshold | Sensitivity | Specificity | Percentage of patients correctly classified | Area under the curve | Odds Ratio |
|---|---|---|---|---|---|
| **>8** | 84.44% | 91.87% | 89.88% | 0.88 (0.82-0.94) | 61.34 (21.82-172.42) |
| **≥11** | 77.78% | 95.93% | 91.07% | 0.87 (0.80-0.93) | 82.60 (26.47-257.72) |
| **>12** | 66.67% | 95.93% | 88.10% | 0.81 (0.74-0.88) | 47.20 (15.89-140.19) |
| **>14** | 57.78% | 97.56% | 86.90% | 0.78 (0.70-0.85) | 54.74 (15.08-198.72) |
